# Air sampling for facility-based respiratory pathogen surveillance in rural Zambia: a prospective comparison with human surveillance

**DOI:** 10.64898/2026.10.28.26366049

**Authors:** Samuel Baron, Mutinta Hamahuwa, Passwell Munachoonga, Pamela Sinywimaanzi, Thomas Aichele, Katherine Fenstermacher, Richard Rothman, Andrew Pekosz, Edgar Simulundu, Shelby L. O’Connor, David H. O’Connor, Chipampe Lombe, Catherine G. Sutcliffe

## Abstract

Respiratory infections are the leading cause of acute disease incidence globally and disproportionately burden low-and middle-income countries where surveillance capacity is limited. While facility-based surveillance remains the standard for respiratory pathogen monitoring in human populations, environmental air sampling offers potential advantages as a non-invasive supplementary method. We conducted a prospective environmental air sampling study nested within ongoing respiratory disease surveillance at Macha Hospital in Southern Province, Zambia, from July 2025 to June 2026. Air samplers were deployed continuously in the outpatient department and three inpatient wards, with filters collected and tested three times weekly. Surveillance in the patient population was conducted year-round with nasopharyngeal samples collected from patients with influenza-like illness. All samples were tested for influenza A/B virus, respiratory syncytial virus (RSV), and SARS-CoV-2 using the Cepheid GeneXpert platform. We analyzed concordance between air and nasopharyngeal samples using percent agreement, Cohen’s Kappa, Spearman correlations, and logistic regression with generalized estimating equations. Over 12 months, 596 air samples and 701 human samples were analyzed. Influenza A virus was the most common pathogen, detected in 38 air samples and 87 nasopharyngeal samples, with most cases detected from July to September 2025. For Influenza A, there was strong same-week concordance between air and nasopharyngeal samples (80.4% overall agreement; 44.4% positive agreement; Cohen’s Kappa = 0.5; Spearman’s rho = 0.57). SARS-CoV-2 was also common, detected in 18 air samples and 16 nasopharyngeal samples, with cases detected sporadically throughout the year. For SARS-CoV-2, there was limited concordance between air and nasopharyngeal samples (68.6% agreement; 23.8% positive agreement; Cohen’s Kappa = 0.2; Spearman’s rho = 0.2). Influenza B virus (1 air and 13 nasopharyngeal samples positive) and RSV (0 air and 4 nasopharyngeal samples positive) circulated sporadically, and unsurprisingly, gave limited concordance between samples. Cycle threshold values for influenza A virus (median for target 1: 36.7 vs. 25.1; p<0.001; median for target 2: 39.5 vs. 28.6; p<0.001) and SARS-CoV-2 (median: 41.3 vs. 27.5; p<0.001) were significantly higher for positive air samples than for nasopharyngeal samples. Implementation challenges were documented, including intermittent power outages and compromised electrical outlets causing sampler shutdowns, as well as study awareness issues that required sensitization activities for both facility staff and patients. Air sampling was feasible and demonstrated promise as a supplementary method for facility-based respiratory pathogen surveillance in rural resource-limited settings. These findings support integrating environmental air sampling alongside surveillance in patient populations as part of a multimodal approach to respiratory pathogen monitoring.

## Introduction

Respiratory tract infections (RTIs) are the leading cause of acute disease incidence and a leading infectious of death globally [1,2]. There were an estimated 12.8 billion episodes of upper RTIs in 2021, and there were an estimated 2.5 million deaths from lower RTIs globally in 2023 [2,3].

Viral infections are a major contributor to the burden of both upper and lower RTIs [2,3], predominantly from seasonal influenza viruses, respiratory syncytial virus (RSV), and human coronaviruses such as SARS-CoV-2 [1,2,4,5].

These viral respiratory infections can cause outbreaks, epidemics, and pandemics [4,6]. For influenza viruses and coronaviruses, this threat stems from their high potential for mutation and gene reassortment which allow for escape from pre-existing immunity [4,7]. These viruses also pose a pandemic threat due to the presence of related viruses in animal reservoirs, most of which are antigenically distinct from human influenza viruses and coronaviruses. For RSV, this is driven by seasonal circulation, waning immunity in vulnerable populations, and increased disease severity in young children and older adults [5,8,9]. Given the high burden of disease and pandemic potential, the global health community has designated respiratory viruses as a high priority for surveillance and response [10].

Traditionally, influenza surveillance has relied on facility-based sentinel surveillance, involving the tabulation of case counts from patients visiting health facilities with a clinical presentation matching a pre-determined case definition for influenza-like illness (ILI) and/or with laboratory-confirmed infection [11,12]. This type of surveillance is resource-intensive, in terms of both human capacity and cost, thus limiting the scale of implementation, particularly in resource-limited settings. In Africa, many countries conduct some level of influenza surveillance and report cases into the Global Influenza Surveillance and Response System (GISRS) [10].

However, the number of years of surveillance and number of surveillance sites in each country vary (e.g., Mozambique reported 72 sentinel sites in 2017 while Zambia reported 6) [13]. In a survey of surveillance programs from 2017, the cost of surveillance ranged from $10,000 to $1.3 million per year; overall funding for these activities in the region had decreased by half since 2011 [13].

Novel, non-case-dependent methods have emerged as supplementary approaches, including wastewater surveillance [15], event-based digital disease surveillance [16–18], and facility-or venue-based air sampling surveillance [19–21], which may provide less resource-intensive options for monitoring respiratory pathogens. There is renewed interest in air sampling as a method for infectious disease surveillance, with most contemporary studies on respiratory pathogen surveillance published in the past decade [22]. Its applications have evolved from environmental air quality monitoring [23,24] and bioaerosol sampling in commercial livestock settings [31,32] to healthcare-associated pathogen detection [21,34,35] and respiratory pathogen surveillance in both healthcare [20,36,37,41–45] and community settings [46–52]. The COVID-19 pandemic accelerated development of these methodologies as the limitations of traditional surveillance became apparent, including delays in case detection as a result of test availability and changes in care-seeking behavior, the effects of severity biases on testing results, and infrastructure gaps for testing in under-resourced healthcare facilities [43,45,49,50,53].

Despite growing evidence and the potential utility of air sampling for resource-limited settings, few studies have been conducted in low- and middle-income countries [45]. The objective of this study was to evaluate the feasibility and utility of air sampling for facility-based respiratory pathogen surveillance in rural Zambia, comparing air sampling with traditional sentinel surveillance.

## Materials and methods

### Study design and setting

This was a prospective facility-based environmental air sampling study nested within ongoing sentinel surveillance for influenza and other respiratory infections. The study was conducted at Macha Hospital in Southern Province, Zambia, a rural facility serving a catchment population of over 150,000 individuals [54]. The area experiences three seasons: a warm rainy season from November to April, a cool dry season from May to August, and a hot dry season from September to October [55]. In collaboration with Macha Research Trust (MRT), an independent research organization [56], the Johns Hopkins Center for Excellence in Influenza Research and Response (JH-CEIRR) established surveillance for respiratory infections at Macha Hospital in 2018 to better understand the epidemiology of respiratory pathogens in this area [55,58]. In 2025, air monitoring was initiated in parallel with surveillance in the patient population.

### Air sampling

In July 2025, Apollo air samplers (Indoor Biotechnologies, Inc., Charlottesville, VA) were continuously deployed in the outpatient department and in the male, female, and pediatric inpatient wards. The Apollo is a quiet, lightweight ambient air sampling device that uses a fan to pull air through an expendable cartridge filter at a high flow rate (>500 L/min) [61]. Samplers were placed to maximize virus detection, with consideration given to wall placement height, air intake and flow, and distance from open windows (S1 Fig). New filters were installed and old filters removed on Monday, Wednesday, and Friday of each week.

In the MRT Clinical Research Laboratory, study staff removed the filter from the plastic frame and transferred it to a 5 mL tube containing Phosphate-Buffered Saline with 0.1% Tween-20 (PBST) solution. The filter was mixed and allowed to soak for 15 minutes to extract sample material into the elution buffer before testing.

### Surveillance in the patient population

The surveillance methods have been previously described [55]. Briefly, all patients visiting the outpatient department were screened for ILI, defined as documented (≥38°C) or self-reported fever in combination with cough or sore throat, with onset or worsening in the past 7 days. An age-stratified (<1, 1–4, 5–15, 16–50, and ≥51 years) sample of outpatients with ILI (target n = 16 per week) was approached for enrollment. Study staff also conducted daily surveillance in the male, female, and pediatric inpatient wards to identify and screen patients admitted for respiratory complaints for ILI. All inpatients with ILI were approached for enrollment. At enrollment, a questionnaire was administered and a nasopharyngeal swab (Copan Diagnostics, Inc., Murrieta, CA) was collected, placed in universal transport media, and transported to the MRT Clinical Research Laboratory.

### Laboratory methods

Nasopharyngeal swabs and air samples were tested at the MRT Clinical Research Laboratory using the GeneXpert Xpress Flu/RSV/SARS-CoV-2 *plus* assay (Cepheid, Sunnyvale, CA) [60]. The assay detects and differentiates influenza A virus (IAV), influenza B virus (IBV), RSV, and SARS-CoV-2. The assay targets the basic polymerase (PB2, referred to as IAV1) and acidic protein (PA, referred to as IAV2) genes of IAV; the nonstructural (NS) protein gene of IBV; and the nucleocapsid genes of RSV A and RSV B; and the nucleocapsid (N2), envelope protein (E), and RNA-dependent RNA polymerase (RdRP) genes of SARS-CoV-2 [61]. Samples were reported to be positive based on the Manufacturer’s algorithm. For each sample, cycle threshold (Ct) values and the qualitative result (positive/negative) were recorded.

### Statistical analysis

The analysis included data from July 14, 2025, to June 29, 2026. Statistical analyses were conducted using RStudio version 5 (Posit PBC, Boston, MA) and Stata version 14 (StataCorp LLC, College Station, TX). For the analysis, samples were considered positive based on the qualitative result reported by Cepheid on the GeneXpert instrument. For each pathogen, location, and type of sampling (air and human), the number of samples collected and testing positive were summarized and heatmaps of weekly positivity were created. For each heatmap of a given location and type, a week was considered positive for a given pathogen if any samples collected in that week were positive for that pathogen. Given the low number of samples per location, results across locations were aggregated for each type of sampling for the remainder of the analysis. Weekly percent positivity (i.e., the proportion of tested air/human samples that were positive in a given week) was calculated for each pathogen and plotted for each type of sampling. Ct values were summarized as median and interquartile range and compared between air and human samples for each pathogen using the Wilcoxon rank-sum test. Spearman’s rank correlation was calculated to compare weekly percent positivity between air and human samples. Concordance between test results for the air and human samples was assessed using several measures, including overall agreement, percent positive agreement, Cohen’s Kappa with bootstrap 95% confidence intervals (CI), and logistic regression with robust standard error estimation. A lagged analysis was also conducted using logistic regression with robust standard error estimation to assess concordance between test results for air samples in week *t* and human samples in week *t+1.* The logistic regression analyses were conducted only for IAV and SARS-CoV-2 due to the limited number of samples testing positive for IBV and RSV.

### Ethical considerations

The study was reviewed and approved by the Institutional Review Boards at the Johns Hopkins School of Medicine (IRB00168163) and Macha Research Trust (E.2018.02), and by the National Health Research Authority (NHRA) in Zambia. Adult participants and the parents or legal guardians of pediatric participants provided written informed consent. Written assent was obtained from children 12 to 15 years of age.

## Results

Between July 14, 2025, and June 29, 2026, 596 air samples were collected (Table 1; S1 File), including 38 (6.4%) positive for IAV, 18 (3.0%) for SARS-CoV-2, and 1 (0.2%) for IBV (none were positive for RSV). Over the same period, 701 human samples were collected, including 87 (12.4%) positive for IAV, 16 (2.3%) positive for SARS-CoV-2, 13 (1.8%) positive for IBV, 4 (0.6%) positive for RSV, and 1 (0.1%) positive for both IAV and SARS-CoV-2.

**Table 1.** Summary of samples collected by type and location in Macha, Zambia, July 2025 to June 2026.

|  | Outpatient<br>Department | Male<br>Ward | Female<br>Ward | Pediatric<br>Ward | Total |
| --- | --- | --- | --- | --- | --- |
| **Air samples** | 149 | 149 | 149 | 149 | 596 |
| SARS-CoV-2 | 5 | 6 | 4 | 3 | 18 |
| Influenza A virus | 7 | 9 | 6 | 16 | 38 |
| Influenza B virus | 0 | 1 | 0 | 0 | 1 |
| Respiratory syncytial virus | 0 | 0 | 0 | 0 | 0 |
| **Human samples** | 656 | 1 | 6 | 38 | 701 |
| SARS-CoV-2 | 16 | 0 | 0 | 0 | 16 |
| Influenza A virus | 71 | 0 | 2 | 14 | 87 |
| Influenza B virus | 13 | 0 | 0 | 0 | 13 |
| Respiratory syncytial virus | 3 | 0 | 1 | 0 | 4 |

### Influenza A virus

IAV-positive samples were observed in both the air and human samples from July to September 2025 and March to May 2026 (Fig 1 and Fig 2). Most positive air samples came from the pediatric ward (n = 16, 42.1%), though positivity was observed in all four locations (Fig 3).

**Fig 1.**
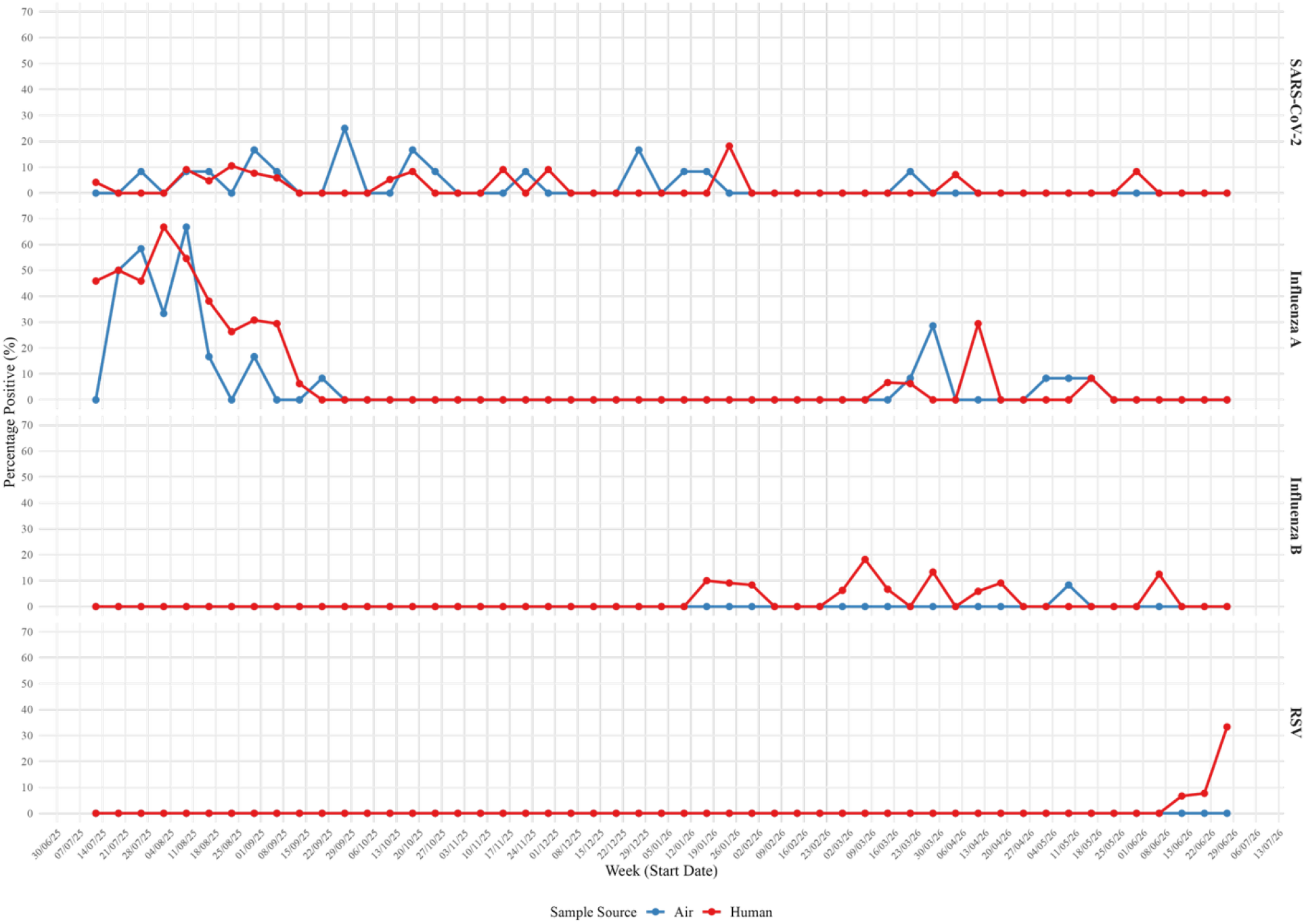
Weekly percent positivity of pathogens by testing method. Time series of weekly proportion of samples testing positive for SARS-CoV-2, influenza A virus, influenza B virus, and respiratory syncytial virus (RSV) in air sampling (blue) and human monitoring (orange), July 2025–June 2026.

**Fig 2.**
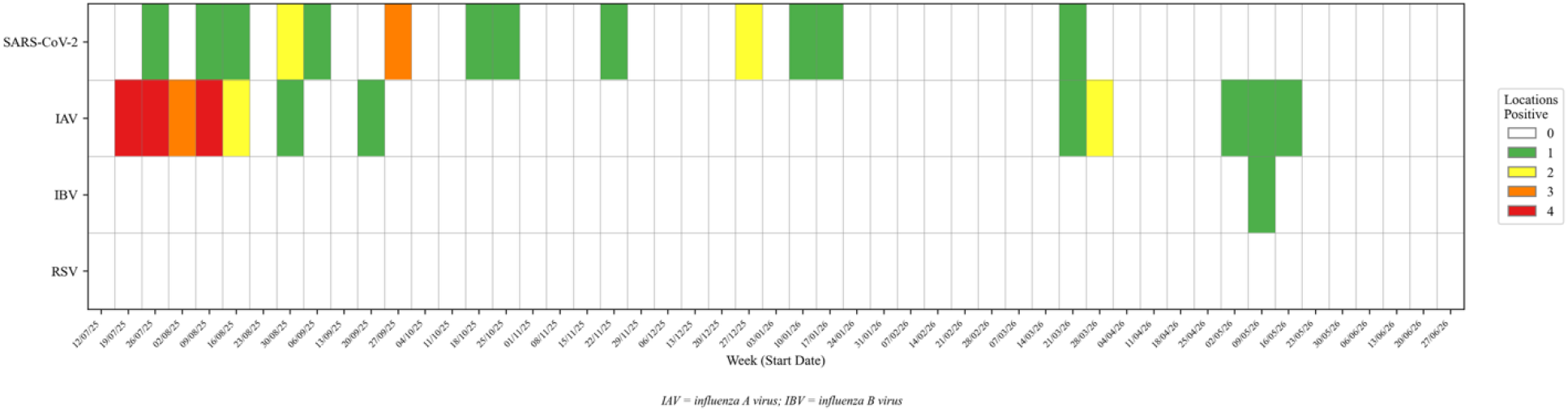
Air sampling positivity by pathogen and date, stratified by number of positive locations. Time series showing the number of hospital locations with positive air samples for SARS-CoV-2, influenza A virus, influenza B virus, and respiratory syncytial virus (RSV), July 2025-June 2026.

**Fig 3.**
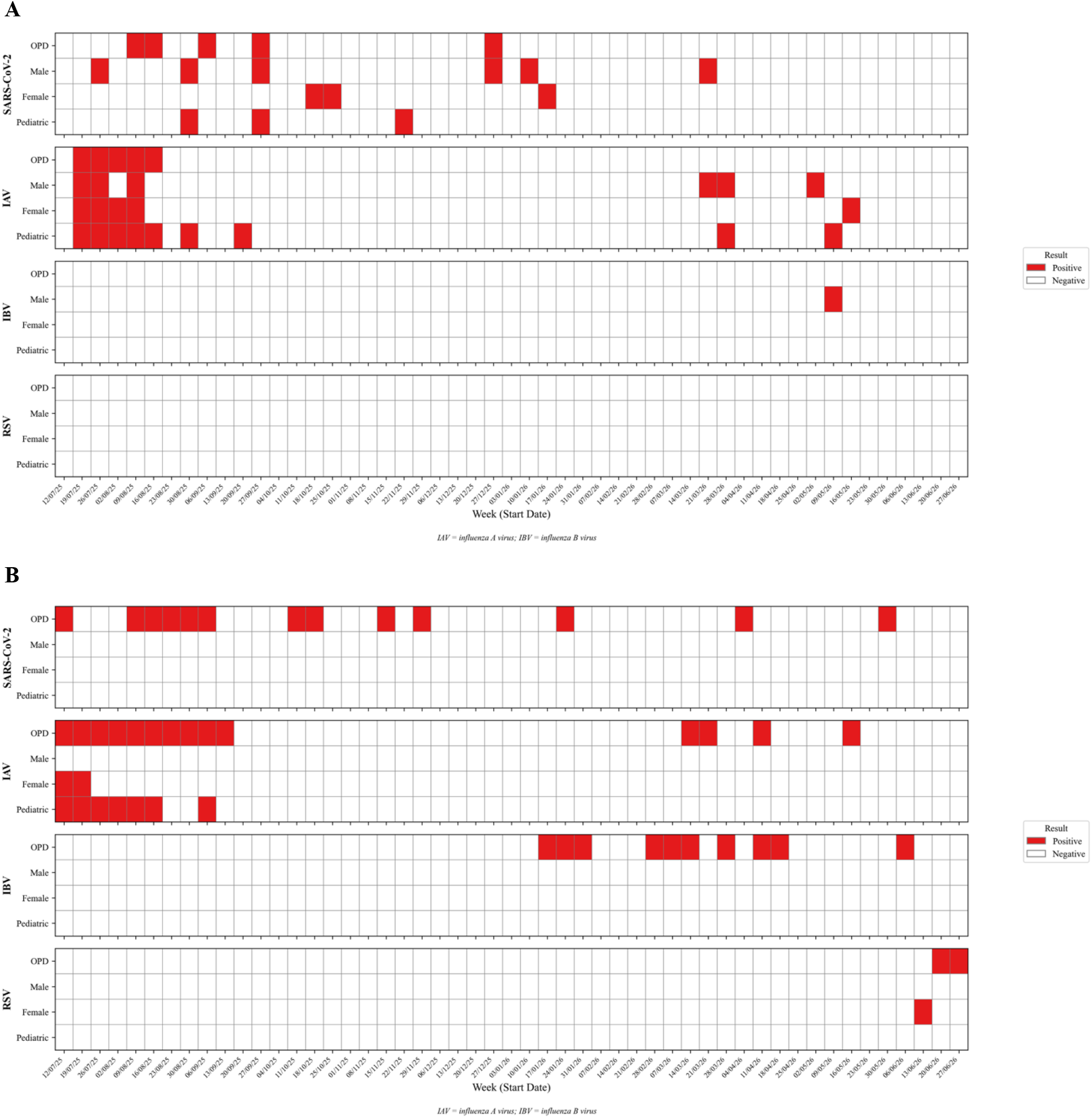
Sampling positivity heatmap by location and pathogen for A) air and B) human monitoring. Heatmap of weekly air sampling positivity across the four hospital locations (outpatient department, pediatric inpatient ward, male inpatient ward, and female inpatient ward) for SARS-CoV-2, influenza A virus, influenza B virus, and respiratory syncytial virus (RSV), July 2025-June 2026.

### SARS-CoV-2

SARS-CoV-2-positive results were observed in both air and human samples in most months of the study period (Fig 1 and Fig 2). Positive results were distributed across wards (range: 3–6 weeks with at least one positive sample for SARS-CoV-2 per ward) (Fig 3).

The median Ct value for SARS-CoV-2-positive air samples was 41.3 (IQR: 39.0, 43.3, p-value <0.001), which was significantly higher than for human samples (median: 27.5, p-value <0.001) (Table 2).

**Table 2.** Cycle threshold (Ct) values for positive samples by pathogen and sample type in Macha, Zambia, July 2025 to June 2026.

| Source |  | SARS-CoV-2 | Influenza A1 | Influenza A2 | Influenza B | RSV |
| --- | --- | --- | --- | --- | --- | --- |
| Air | N | 18 | 38 | 24 | 1 | — |
|  | Ct | 41.3 [39.0, 43.2] | 36.7 [35.9, 37.8] | 39.5 [38.2, 41.1] | 37.8 [37.8, 37.8] | — |
| Human | N | 13 | 87 | 87 | 13 | 4 |
|  | Ct | 27.5 [22.2, 30.5] | 25.1 [22.1, 28.9] | 28.6 [25.0, 32.0] | 23.6 [22.6, 26.3] | 26.9 [26.2, 28.0] |
| P-value |  | <0.001 | <0.001 | <0.001 | 0.14 | — |

| Source |  | SARS-CoV-2 | Influenza A1 | Influenza A2 | Influenza B | RSV |
| --- | --- | --- | --- | --- | --- | --- |

Same-week concordance between air and human positive was limited for SARS-CoV-2, as demonstrated by overall agreement (68.6%), percent positive agreement (23.8%), Cohen’s Kappa (0.2; bootstrap 95% CI: −0.1 to 0.5), Spearman’s rho (0.2; p = 0.27), and logistic regression (OR; 2.3; 95% CI: 0.6, 9.2) (Table 3, Fig 4).

**Fig 4.**
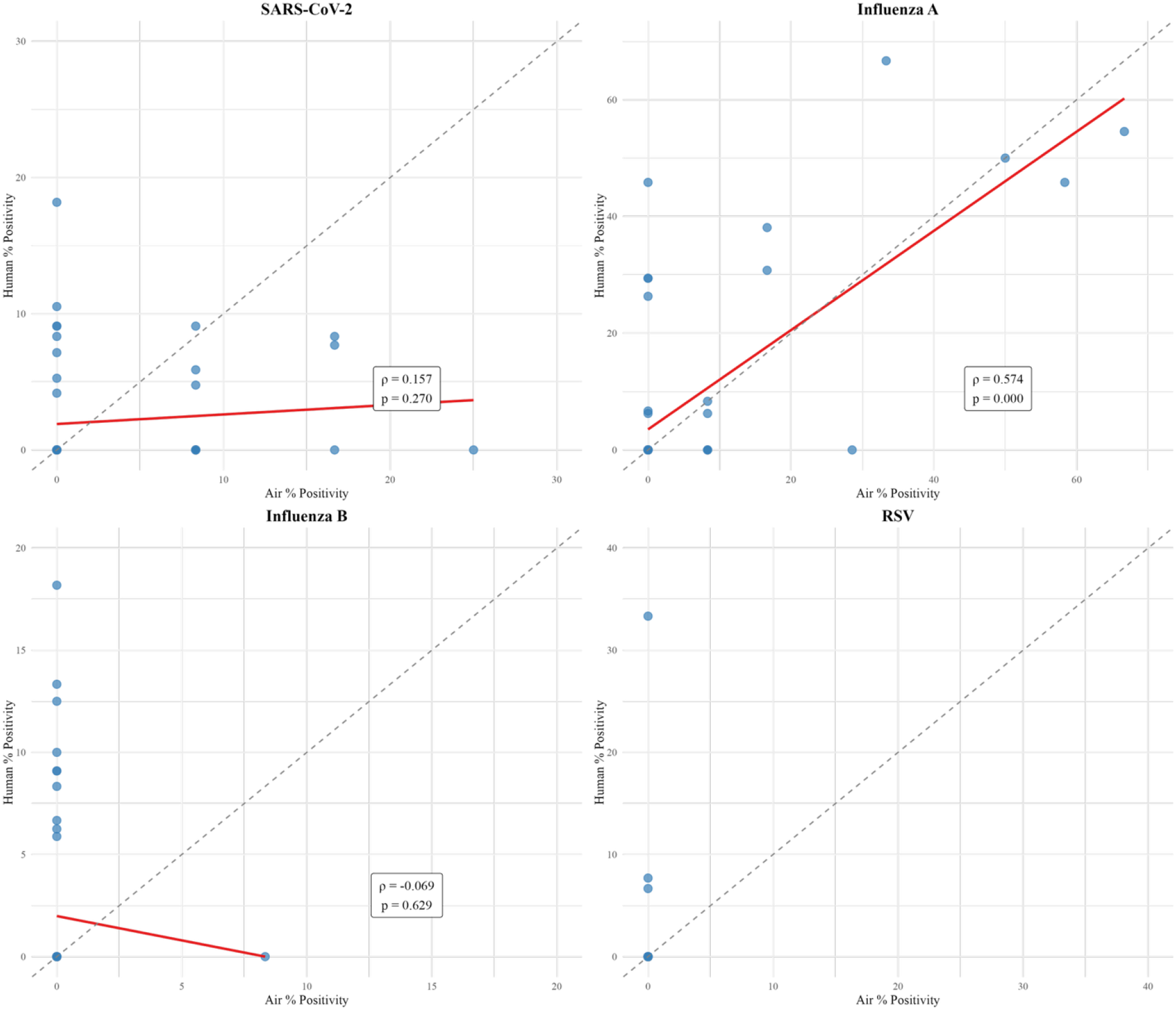
Correlation of percent positivity between sampling methods. Scatterplots of weekly percent positivity in air versus human samples for SARS-CoV-2, influenza A virus, influenza B virus, and respiratory syncytial virus (RSV) with Spearman’s rho and p-values.

**Table 3.** Same-week concordance between air and human pathogen detection in Macha, Zambia, July 2025 to June 2026.

| Category | Measure | SARS-CoV-2 | Influenza A virus | Influenza B virus | RSV |
| --- | --- | --- | --- | --- | --- |
| Weekly Agreement Summary | Human +/Air+ | 5 (9.8%) | 8 (15.7%) | 0 (0.0%) | 0 (0.0%) |
|  | Human-/Air- | 30 (58.8%) | 33 (64.7%) | 40 (78.4%) | 48 (94.1%) |
|  | Human+/Air- | 8 (15.7%) | 6 (11.8%) | 10 (19.6%) | 3 (5.9%) |
|  | Human-/Air+ | 8 (15.7%) | 4 (7.8%) | 1 (2.0%) | 0 (0.0%) |
| Agreement Metrics | Overall Agreement | 68.6% | 80.4% | 78.4% | 94.1% |
|  | Percent Positive Agreement | 23.8% | 44.4% | 0% | 0% |
|  | Cohen's Kappa (95% CI)<sup>a</sup> | 0.2 (-0.1–0.5) | 0.5 (0.2–0.7) | N/A | N/A |
| Association Measure | Odds Ratio (95% CI)<sup>b</sup> | 2.3 (0.6–9.2)<br>p=0.22 | 11.0 (2.5–48.4)<br>p=0.002 | N/A | N/A |

OR = Odds Ratio; CI = Confidence Interval
Green = concordant results; Red = discordant results
N/A: Cohen's Kappa and Odds Ratio were not calculated for influenza B virus and respiratory syncytial virus (RSV) due to insufficient positive samples or quasi-complete separation
<sup>a</sup> Bootstrap 95% confidence intervals (CI) reported for Kappa values
<sup>b</sup> GLM + Robust SE: Week-level analysis (N=51 week-pairs) with heteroscedasticity-consistent standard errors

In the lagged-week analysis, there was no statistically significant temporal association between air sampling in a given week and human sampling in the following week (OR = 2.7; 95% CI: 0.7, 10.7) (Table 4).

**Table 4.** Lagged-week temporal association between air sampling (week *t*) and human sampling (week *t*+1)

| Category | Measure | SARS-CoV-2 | Influenza A virus |
| --- | --- | --- | --- |
| **Weekly Agreement Summary**<br>**(Air week $t \rightarrow$ Human week $t+1$)** | Air+ → Human+ | 5 (10.0%) | 7 (14.0%) |
| | Air- → Human- | 30 (60.0%) | 32 (64.0%) |
| | Air+ → Human- | 8 (16.0%) | 5 (10.0%) |
| | Air- → Human+ | 7 (14.0%) | 6 (12.0%) |
| **Association Measure** | Odds Ratio <sup>a</sup> (95% CI) | 2.7 (0.7–10.7) | 7.5 (1.8–31.6) |
|  | P-value | 0.16 | 0.006 |

OR = Odds Ratio; CI = Confidence Interval
Green = concordant results; Red = discordant results
<sup>a</sup> GLM + Robust SE: Week-level analysis (N=50 week-pairs) with heteroscedasticity-consistent standard errors

### Influenza B virus

Only 1 air sample and 13 human samples tested positive for IBV, primarily from January to June 2026 (Fig 1 and Fig 2). The Ct value for the IBV-positive air sample was 37.8 compared to a median of 23.6 for the human samples (p-value=0.14) (Table 2). Overall agreement was high (78.4%) given the low number of cases; percent positive agreement was 0%.

### RSV

No air samples and 4 human samples tested positive for RSV in June 2026 (Fig 1 and Fig 2). Overall agreement was high (94.1%), but given the low number of cases, percent positive agreement was 0%.

### Implementation challenges

The project faced several practical challenges to implementation. The most common issue encountered concerned the power supply at Macha Hospital. The outlets powering the air samplers, particularly the one in the outpatient department, were loose, allowing the power cords to be easily dislodged. In addition, patients and facility staff would occasionally unplug the samplers for personal use, and facility-wide power outages occurred. This resulted in an unquantified loss of sampling time for some sampling periods. A second issue was that patients and staff occasionally moved the samplers, resulting in a change of intake position, which could have decreased the quantity of room air directed through the sampler. To mitigate these issues, facility staff were regularly briefed on the function of the samplers, and signs were placed above the samplers with instructions not to unplug or move the samplers. In addition, power strips were provided so that patients and staff would have outlets for other uses. A third recurring issue was the intake of small insects and dust, particularly during the dry season, into the air samplers and onto the filters (S2 Fig), which could have affected sample integrity and handling. Fourth, when air sampling was initiated, some patients expressed concerns about the presence of the devices (e.g., they thought they were cameras). Study staff addressed these concerns by briefing patients and their families on the purpose of the samplers.

## Discussion

In this study comparing respiratory virus detection by facility-based monitoring of the air and patient population in rural Zambia, we found that air sampling was feasible to implement and identified similar periods of virus circulation as surveillance in the patient population, particularly for IAV and SARS-CoV-2. These findings support the utility of air sampling as a complementary method for assessing respiratory pathogen circulation in low-resource healthcare settings.

This study compared two methods that monitor respiratory viruses in different ways. Human sampling reflects virus replication in enrolled symptomatic patients, with detection impacted by the volume and frequency of samples collected. Air sampling reflects airborne viral circulation in the sampled clinical environment, which includes symptomatic patients (who may have been directly sampled), but also other patients, healthcare workers, and visitors. Detection through air sampling will be impacted by sampler placement and proximity to patients and other people.

Given these differing methodologies, concordance between the two reflects their ability to detect viral circulation in their sampled populations at the same time, and some level of discordance may be expected.

In this study, the highest level of agreement and concordance between the two methods was observed for IAV. In Macha, IAV is typically seasonal [54], resulting in concentrated periods of high positivity among patients. During the study period, this was observed from July to September 2025, with up to 67% of patients testing positive in a week, followed by a decline and then low levels of detection from March to May 2026. This concentrated circulation in 2025 likely increased the likelihood that both surveillance modalities would detect the same underlying period of disease transmission.

In contrast to IAV, lower agreement was observed for SARS-CoV-2, IBV, and RSV, which were only detected sporadically and at low levels during the study period. By the time of this study, SARS-CoV-2 transmission in Macha had transitioned from pandemic waves to sustained low-level transmission [68]. This sporadic transmission pattern likely contributed to the observed discordance between air and nasopharyngeal samples, as detection for air sampling would be more likely impacted by the placement of the samplers near infected patients, and detection in the patient population would be more likely impacted by which patients were selected for enrollment. Similar results, with stronger associations for IAV than SARS-CoV-2, were observed in a comparison of air and human monitoring in schools in the United States in 2022-2023 [36, 69]. These results are also consistent with a study from an emergency department that found that the number of patients positive for SARS-CoV-2, influenza virus or RSV was significantly higher during periods when the sampler was positive than negative, suggesting that sufficient numbers of cases need to be present for a given virus to be detected in the air.

The air samples produced consistently higher Ct values compared to nasopharyngeal swabs for all pathogens. The high Ct values in this study are consistent with those from an air sampling study in congregate settings in the United States [50]. This suggests that air samples capture less viral material, consistent with the dilution of respiratory secretions into ambient air compared to direct sampling at the site of viral replication in patients. In this study, air was sampled continuously over two or three days and then tested. In one study from the United States, similar results and Ct values were obtained from continuous daily and weekly sampling [50], thereby extending the utility of air sampling for surveillance in settings where resources can only sustain weekly sampling.

The physical environment at Macha Hospital differs from health facilities in high-income country settings. At Macha Hospital, the windows and doors in the outpatient department and the wards are typically open, providing continuous natural ventilation. This contrasts with the sealed, mechanically ventilated environments in health facilities in high-income settings where some prior air sampling studies have been conducted [22,33], and is more similar to airports and communal settings where other air sampling studies have been conducted [50,52,69]. Our positive results in this open-air environment demonstrate that air sampling methods remain viable in such contexts, an important finding for generalizing this approach to similar health facilities in resource-limited settings. We documented practical challenges unique to resource-limited settings, including intermittent power outages and compromised electrical outlets that caused sampler shutdowns. These could be mitigated by decoupling the samplers from an unreliable power grid through the use of batteries or solar power. Several study awareness issues were also encountered that required sensitization activities for both facility staff and patients, highlighting important considerations when designing environmental surveillance programs for resource-limited settings.

Facility-based surveillance in patient populations and air sampling provide different but complementary information about respiratory pathogen circulation. While monitoring patient populations has been the standard for surveillance and produces rich data on both cases and pathogens, it is resource-intensive, thus limiting its scope. Air sampling can provide information on pathogens present in a location, and as it can be implemented with fewer resources, surveillance platforms that include air sampling may be able to expand across a broader set of facilities and populations. However, air sampling cannot provide clinical and demographic data on the cases and may encounter limitations in the quantity of viral material available for downstream analyses such as sequencing [15,19,37]. Integrating these different modalities into surveillance platforms may help health systems more comprehensively characterize pathogen circulation.

Several limitations should be acknowledged. First, air sampling was initiated partway through the influenza season in 2025, precluding assessment of concordance at the onset of circulation. Second, the relatively small sample size (51 study weeks) and number of cases limited statistical power, particularly for IBV and RSV. Insufficient sample positivity at the ward level precluded ward-specific analyses, necessitating aggregation of results across locations. Consequently, the concordance measures reflect the status of the facility as a whole rather than individual wards.

Third, one air sampler was deployed in each location, and in these open clinical spaces, detection likely depended on the transient proximity of infected patients, visitors, or staff to the sampler during the collection period. As such, the results from the air samplers may reflect the immediate microenvironment at their location rather than the ward as a whole. Fourth, viral concentrations per unit volume of air sampled could not be estimated as the exact air volume sampled was not available and the Xpert assay is semi-quantitative, providing Ct values instead of viral load.

Lastly, this study was conducted at one hospital in rural Zambia with detection of four viruses, and the results may not be generalizable to all facilities in rural areas in the region or for all viral pathogens. Future research should prioritize evaluation over a broader range of settings and pathogens.

## Conclusions

This study provides evidence that air sampling is a viable and feasible supplementary method for facility-based respiratory pathogen surveillance in rural resource-limited settings. Sampling the air and patient population provided similar inferences about pathogen circulation, demonstrating strong concordance for viruses circulating seasonally and lower concordance for viruses circulating at lower levels or sporadically. These findings support integrating environmental air sampling alongside surveillance in patient populations as part of a multimodal approach to respiratory pathogen monitoring.

## Data Availability

Summary testing data are provided in the Supplementary Material (S1 File). Participant data used to support the findings of this study are governed by the regulations of the Government of Zambia and are not publicly available for legal and ethical reasons. All investigators interested in the participant data are required to submit a written, reasonable request to the corresponding author Dr. Catherine Sutcliffe or the National Health Research Authority.

## Acknowledgments

The authors thank the study participants, the study team at Macha Research Trust, and the staff at Macha Hospital.

