## Supplemental Material for "Air sampling for facility-based respiratory pathogen surveillance in rural Zambia: a prospective comparison with human surveillance"

#### Supplemental Figures

**S1 Figure. Air sampler locations in Macha Hospital.** Photographs showing placement of Apollo air samplers in the A) outpatient department, B) male ward, C) female ward, and D) pediatric ward. Blue boxes represent hospital beds, orange represents benches, and brown represents physical obstructions like walls.

**S2 Figure. Images of the A) air sampler and B) filter during the dry season in July 2026**

#### Supplemental Files

**S1 File. Weekly test results for air and human sampling (excel file)**

**S1 Figure. Air sampler locations in Macha Hospital.** Photographs showing placement of Apollo air samplers in the A) outpatient department, B) male ward, C) female ward, and D) pediatric ward. Blue boxes represent hospital beds, orange represents benches, and brown represents physical obstructions like walls.

### A. Outpatient Department

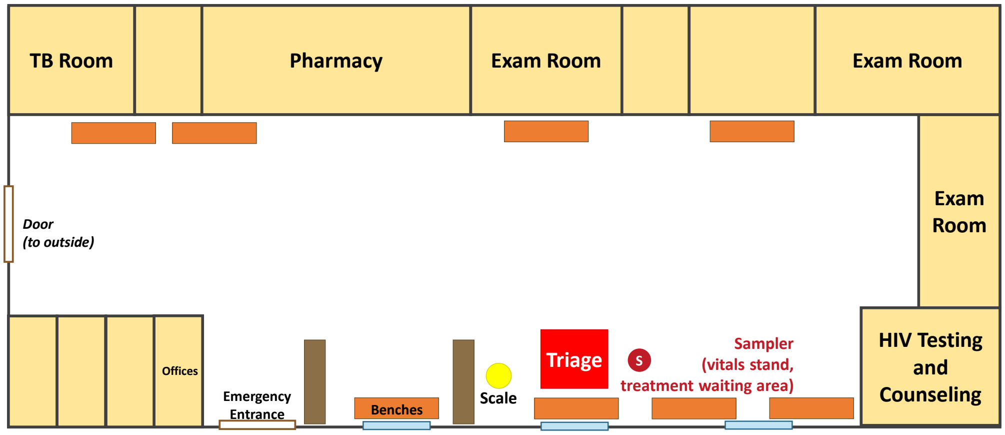


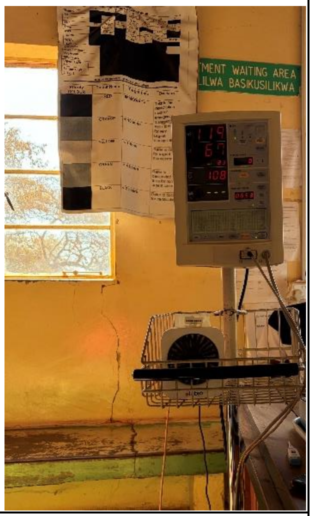

*Layout showing examination rooms along top, central waiting area with benches (orange), triage station (red), and scale (yellow dot). Sampler positioned on vitals stand in treatment waiting area. Blue boxes along the wall are windows, brown outlined boxes are doorways, and brown solid boxes represent internal partitions.*

### B. Male Ward

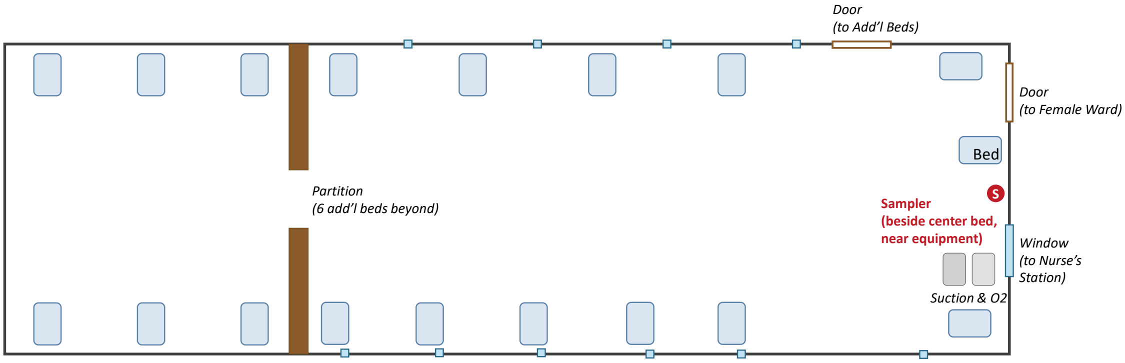


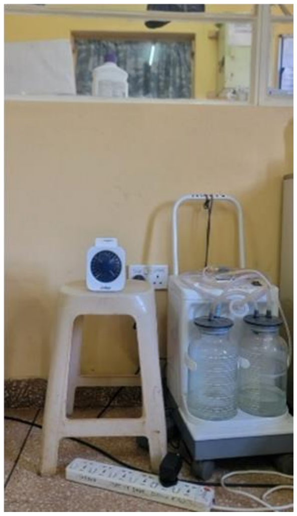

*Layout shows beds along both walls with equipment area and individual bed on right side near sampler. Brown partition on left (walls stop short of the ceiling) separates six additional beds beyond.*

### C. Female Ward

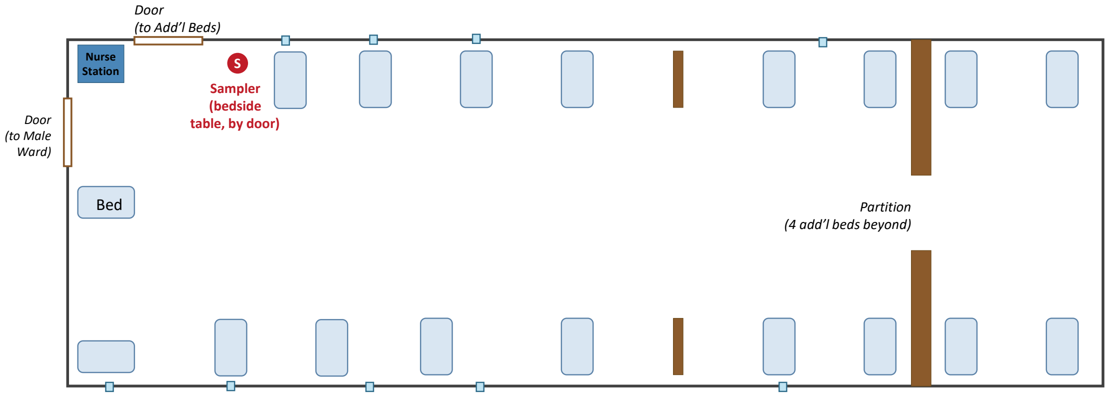


*Layout showing beds along both walls with sampler on a table beside a bed and next to a doorway. Brown wall sections indicate internal partitions, including the end partition (walls stop short of the ceiling) that separates four additional beds beyond.*

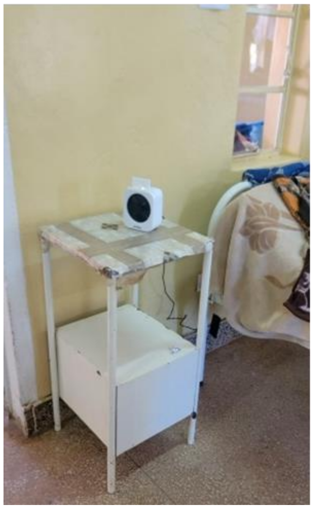

### D. Pediatric Ward

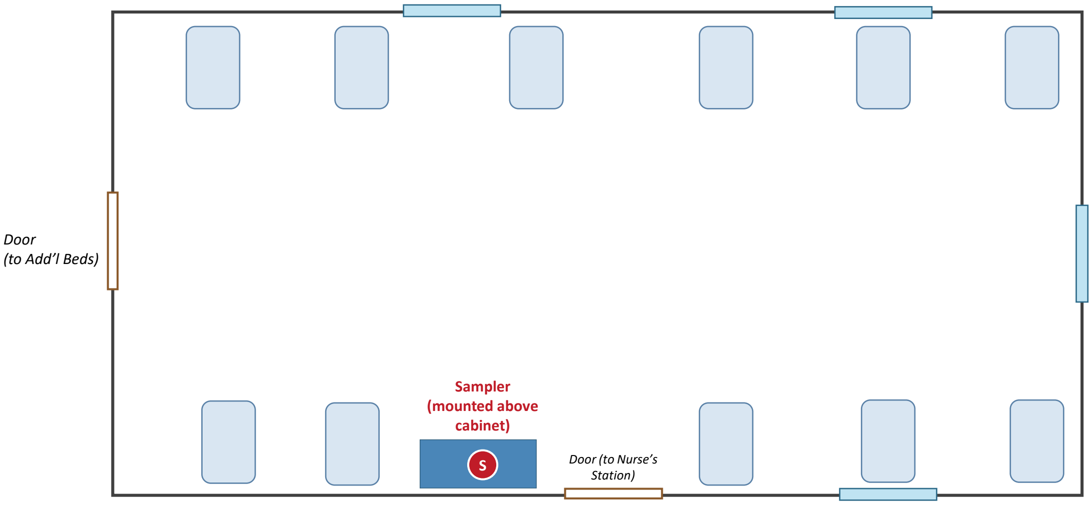


*Layout shows a single room with cots arranged around perimeter. Cabinet next to exit door with sampler mounted above.*

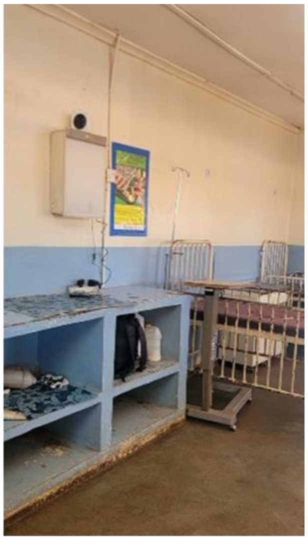

**S2 Figure. Images of the A) air sampler and B) filter during the dry season in July 2026**

**A.** **B.**

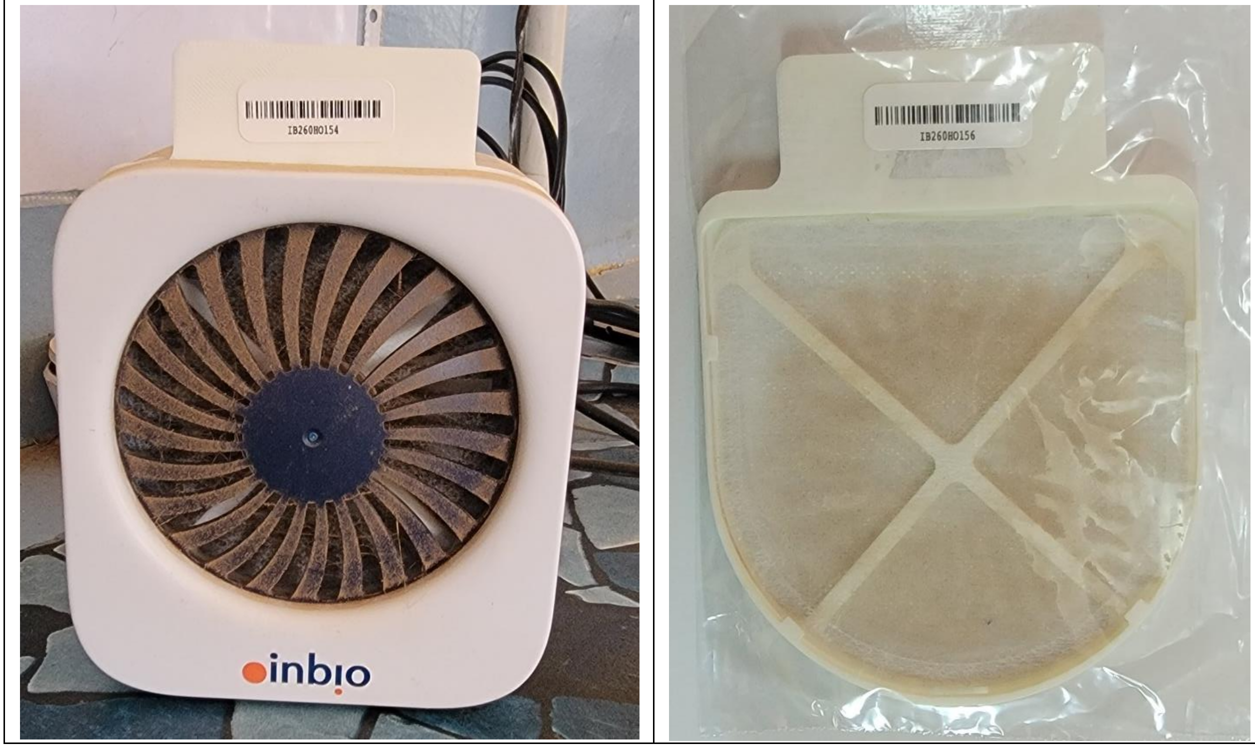
